# Transdiagnostic connectome-based prediction of craving

**DOI:** 10.1101/2021.05.21.21257620

**Authors:** Kathleen A Garrison, Rajita Sinha, Marc N. Potenza, Siyuan Gao, Qinghao Liang, Cheryl Lacadie, Dustin Scheinost

## Abstract

Craving is a central construct in the study of motivation and human behavior and is also a clinical symptom of substance and non-substance-related addictive disorders. Thus, craving represents a target for transdiagnostic modeling. We applied connectome-based predictive modeling (CPM) to functional connectivity data in a large (N=274) transdiagnostic sample of individuals with and without substance-use-related conditions, to predict self-reported craving. CPM is a machine-learning approach used to identify neural ‘signatures’ in functional connectivity data related to a specific phenotype. Functional connectomes were derived from three guided imagery conditions of personalized appetitive, stress, and neutral-relaxing experiences. Craving was rated before and after each imagery condition. CPM successfully predicted craving, thereby identifying a transdiagnostic ‘craving network’ comprised primarily of the posterior cingulate cortex, hippocampus, visual cortex, and primary sensory areas. Findings suggest that craving may be associated with difficulties directing attention away from internal self-related processing, represented in the default mode network.

## Introduction

Craving –the strong desire or urge to engage in motivated action such as use of a substance ^1^ –is central to most models of addiction, ^2^ and is a common phenomenological feature or symptom of addictive disorders that predicts future substance use and treatment outcomes. Exposure to substance cues is associated with the subjective experience of craving, ^3,4^ and individuals with substance use disorders (SUDs) often report craving as a main reason for continued use and relapse. ^5^ Craving has been directly associated with substance use and relapse in individuals with addictions, ^6-8^ and momentary variations in craving have been associated with specific instances of substance use and relapse. ^9^ Moreover, self-reported craving has been included as a testing paradigm for one essential construct –expectancy/reward prediction error –considered by some as ‘primary’ to the understanding of addictive behavior. ^10^

Despite considerable research attention, the clinical significance of craving has been debated, ^11^ particularly the inclusion of craving as a diagnostic criterion for all SUDs, ^12^ due in part to the heterogeneity of craving across different SUDs. ^3,13^ Support for the clinical relevance of craving is bolstered by neuroimaging meta-analyses showing similar neural substrates underlying substance cue-reactivity and craving across SUDs. ^14,15^ However, the included studies typically rely on explanatory analyses to identify neuroimaging measures related to craving measures, which often do not generalize to novel individuals or demonstrate adequate clinical utility. ^16^ There is very limited *predictive* research on brain-phenotype associations, including with craving, across SUDs. Moreover, behavior motivated by reward or appetite, mediated by ‘wanting,’ is experienced by individuals with and without SUDs (e.g., food craving, non-disordered substance use), with addictive behavior motivated by excessive ‘wanting’ of drugs, or drug craving.^17^ Therefore, craving represents a target for transdiagnostic modeling including comparator samples.

Connectome-based predictive modeling (CPM) is a well-validated machine learning approach for generating predictive models of brain-phenotype associations from whole-brain functional-connectivity data, or ‘connectomes.’ ^18^ In contrast to explanatory analyses involving correlations or regression, machine learning models, like CPM, protect against overfitting by being defined and validated in independent samples, leading to more accurate effect sizes and increasing the generalizability of findings. ^19,20^ CPM is also data-driven, and is therefore used to identify neural ‘signatures’ in functional-connectivity data related to a specific phenotype. ^18,21^ CPM has been used to predict cognitive and clinical phenotypes, ^22-25^ and, more recently, has been used to predict treatment outcomes in SUDs. ^26,27^

This study aimed to identify a transdiagnostic ‘craving network’ by applying CPM to functional connectivity data in individuals with and without substance-use-related conditions, to predict a continuous measure of self-reported craving. Functional magnetic resonance imaging (fMRI) data were collected during three types of personalized guided imagery conditions comprising an appetitive cue (favorite food and preferred drug) or stress cue as compared to a control neutral-relaxing cue. Participants rated their craving before and after each imagery condition. For each imagery condition, task connectomes were constructed, and CPM was used to combine task connectomes for the prediction of craving. Based on previous work implicating the default mode network (DMN) in internally-directed attention and self-referential thought processes related to craving, ^28^ it was hypothesized that functional connectivity in the DMN would predict individual differences in self-reported craving.

## Results

In a transdiagnostic sample pooled from several published studies, 274 participants underwent a well-validated personalized guided imagery task during fMRI. Participants included those with substance-use-related conditions, including alcohol use disorder (n=38), cocaine use disorder (n=28), obesity (body mass index > 30 kg/m^2^; n=19), and adolescents with prenatal cocaine exposure (n=41); as well as controls (n=148), including adult controls (n=131) and adolescents without prenatal cocaine exposure (n=17). During fMRI, participants were presented with six personalized guided imagery scripts: two appetitive (favorite food, preferred drug –alcohol, cocaine), two stress, and two neutral-relaxing scripts. Before and after each imagery trial, participants rated their craving on a verbal analog scale from 1 to 10. Craving ratings indicated their “desire to [use substance/food] at that moment,” thus providing concurrent neural and subjective responses per trial.

Independently, for each imagery condition, fMRI data were processed with a validated pipeline and parcellated into 268 nodes using a whole-brain, functional atlas defined previously in a separate sample. ^21^ Next, the mean time courses of each node pair were correlated and correlation coefficients were Fisher transformed, generating three connectivity matrices per participant (i.e., each participant had an appetitive, stress, and neutral-relaxing connectome). These connectomes were subsequently used to predict the individuals’ subjective craving responses.

For predictive modeling, a multidimensional approach was employed, which combines multiple connectomes and behavioral measures into a single predictive modeling framework. ^25^ A principal components analysis (PCA) was used to summarize craving in a data-driven manner for predictive modeling with CPM. To maintain separate train and test groups, for each iteration, only the training datasets were used as inputs to the PCA, and these PCA coefficients were applied to the test dataset. To generate predictive models of craving, all task connectomes were combined into a single predictive model using ridge regression. ^29^ Using multiple connectomes improves predictive modeling and facilitates a more holistic characterization of brain-behavior associations. ^29,30^ Ten-fold cross-validation was used to train transdiagnostic models of craving. In addition, leave-one-group-out cross validation was used to test whether prediction results were driven by group differences in prediction performance from including both controls and individuals with alcohol or cocaine use disorder, obesity, or adolescents with prenatal cocaine exposure in a single model. Model performance was evaluated with a cross-validated coefficient of determination, labeled q^2^, and permutation testing.

### Self-report craving

Craving ratings for the substance-use-related group and the control group are presented in Figure 1. For all conditions, the substance-use-related group reported higher craving compared to the control group (appetitive: t=3.32, p=0.001, df= 296.17; stress: t=4.00, p<0.001, df= 293.07; neutral-relaxing: t=3.49, p<0.001, df= 278.71). In the appetitive and stress conditions, craving ratings after the task were greater than craving ratings before the task (appetitive: t=6.00, p<0.001, df= 257.05; stress: t=2.65, p=0.009, df=261.81). There were no significant group-by-time interactions. All models included gender and age as covariates. Craving ratings from the substance-use-related group exhibited greater variance than craving ratings from the control group (Bartlett c^2^= 53.3, p<0.001, df=3). Finally, craving ratings were highly correlated within and across imagery conditions (r’s>0.86, p’s<0.001, df=272). These correlations were similar for the substance-use-related (r’s>0.85, p’s<0.001, df=129) and control groups (r’s>0.86, p’s<0.001, df=143), independently.

**Figure 1.**
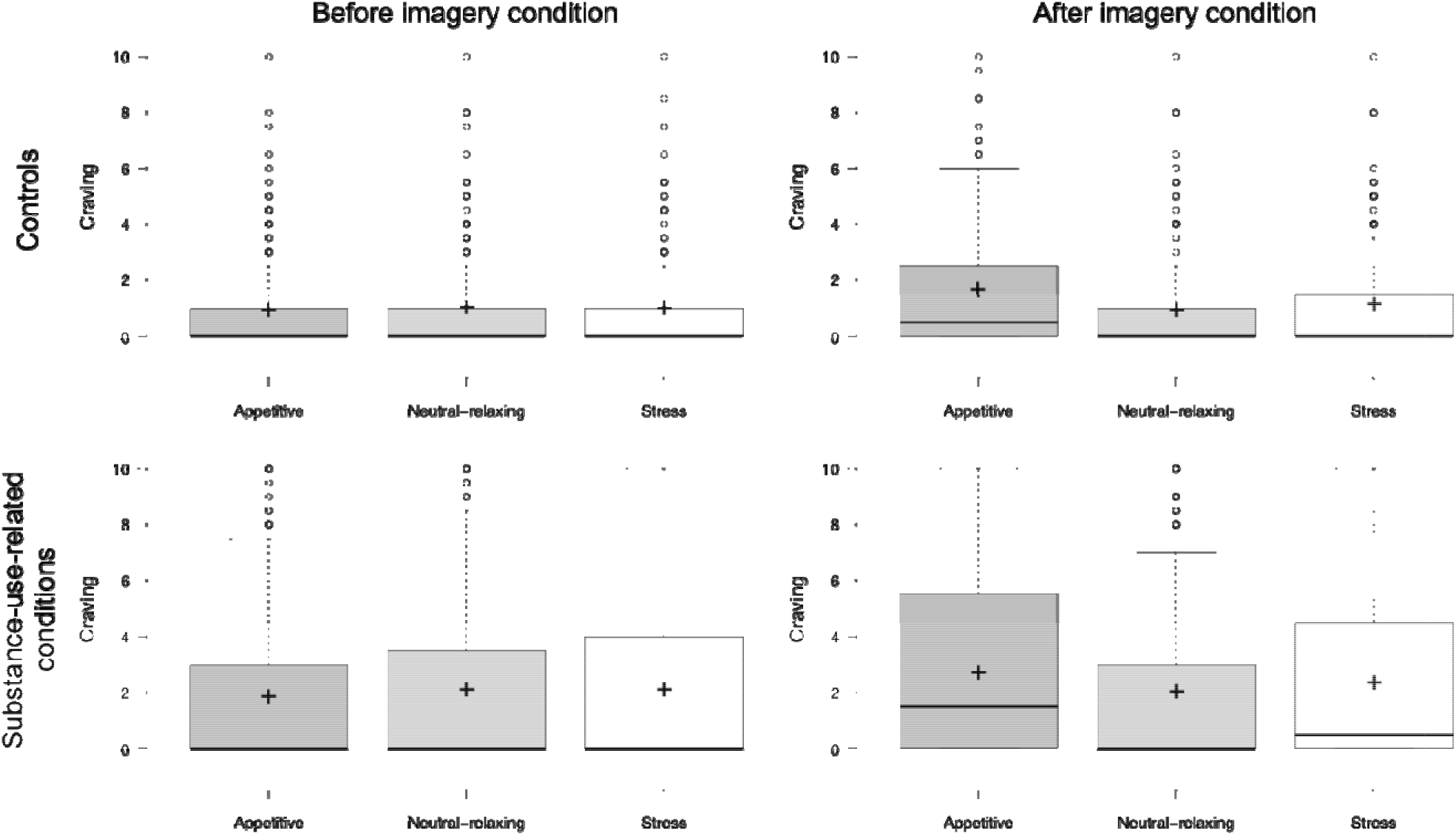
Self-reported craving. Box plots for self-reported craving ratings (verbal analog scale from 1-10) before and after each imagery condition (appetitive, neutral-relaxing, stress) for controls (top) and individuals with substance use related conditions (bottom).

### Transdiagnostic prediction of craving

In a transdiagnostic manner, the overall CPM model successfully predicted self-reported craving as defined by the 1^st^ principal component derived from all craving ratings (r=0.41, ρ=0.37, q^2^=0.14, MSE=34.22, p<0.001, permutation testing 1,000 iterations, 1-tailed; Figure 2). Follow-up comparisons adjusting for head motion (r=0.40, ρ=0.36, q^2^=0.14, MSE=34.28), age (r=0.24, ρ=0.24, q^2^=0.05, MSE=37.88), heart rate (r=0.41, ρ=0.37, q^2^=0.14, MSE=34.16), anxiety ratings (r=0.40, ρ=0.37, q^2^=0.13, MSE=34.62), group (r=0.38, ρ=0.36, q^2^=0.13, MSE=34.73), smoking status (r=0.41, ρ=0.38, q^2^=0.14, MSE=34.08) and sex (r=0.41, ρ=0.37, q^2^=0.14, MSE=34.12) demonstrated similar prediction performances. Each of the six craving ratings (i.e., before and after each imagery condition) contributed approximately equally to the 1^st^ principal component (Figure S2). As such, results were similar when predicting the mean craving, rather than using the 1^st^ principal component (Table S1).

**Figure 2.**
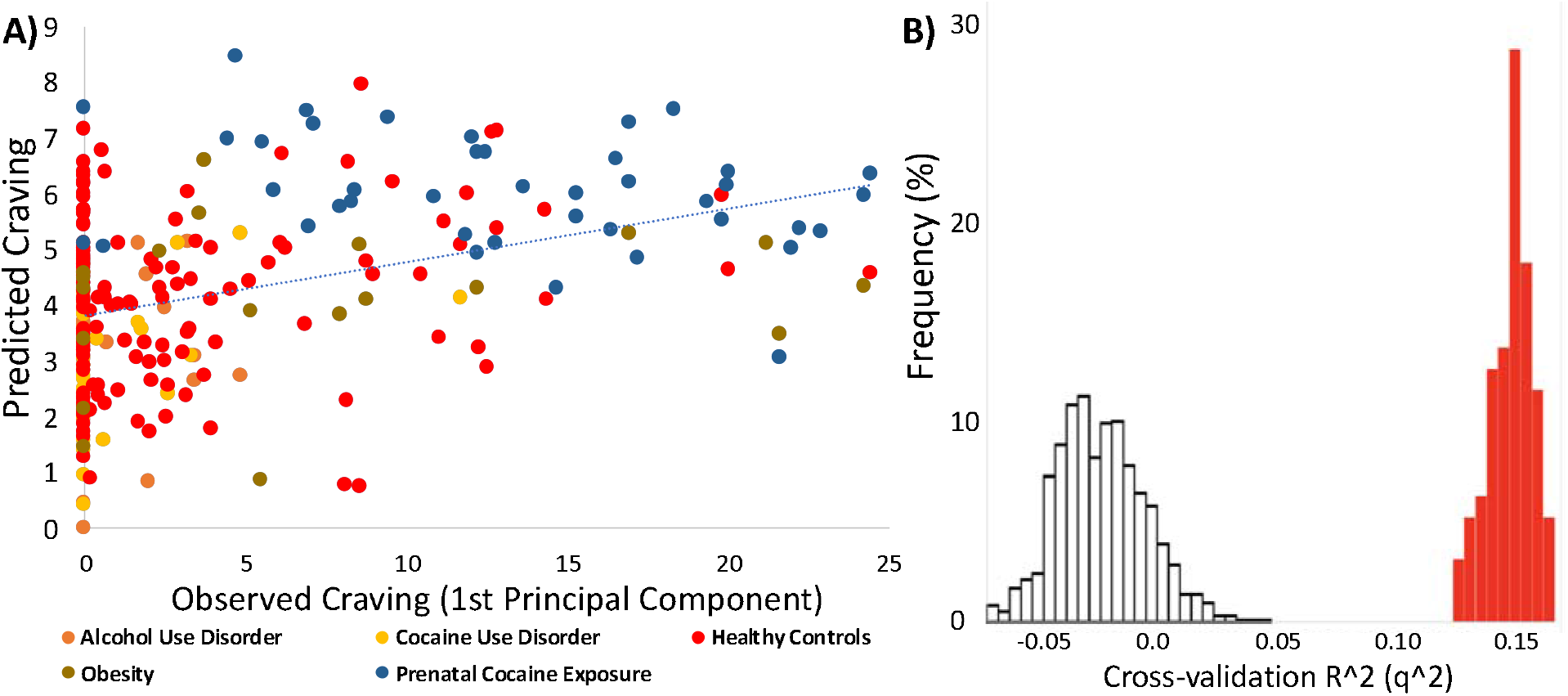
Connectome-based predictive model performance for transdiagnostic prediction of craving. (A) Scatter plot of the observed 1^st^ principal component of the craving data on the x-axis and the predicted principal component generated using CPM on the y-axis. Each group of individuals is colored independently. (B) Histogram of the model performance, indexed by q^2^, across 100 iterations of the actual data (red) and 1,000 iterations of randomly permuted data (white).

While results did not appear driven by better prediction in one group (Figure 2), this was explicitly tested by training solely on either the substance-use-related group or control group, and testing on the other, as these groups differed in craving ratings. CPM was able to predict craving in the substance-use-related group from models trained in the control group (r=0.28, p=0.001, df=143) and vice versa (r=0.19, p=0.02, df=127), showing that prediction performance was high regardless of the training data.

Next, potential differences in prediction performance between craving rated before and after the imagery conditions were tested (Table S1). Notably, prediction of craving measured before the imagery conditions was significantly better than prediction of craving measured after the imagery conditions when using either the 1^st^ principal component (z=3.43, p<0.001) or the mean craving (z=2.14, p=0.032).

Finally, we investigated whether the multidimensional approach of combining connectomes and craving ratings from multiple imagery conditions produced better predictions over using a single imagery condition and craving rating (Table S2). In all cases, prediction using a single imagery condition and craving rating was significantly worse than the multidimensional model (appetitive: z=10.85, p<0.001; stress: z=10.59, p<0.001; neutral-relaxing: z=4.50, p<0.001).

### Anatomical and task contribution to craving prediction

In line with previous CPM results, the identified ‘craving network’ was complex with contributions from every node and canonical brain network. Overall, the top 5% of nodes contributing to craving prediction were located in the posterior cingulate cortex (PCC), hippocampus, visual cortex, and primary sensory areas (Figure 3). Similarly, at the network level, the DMN, motor-sensory, visual, and subcortical networks were the most prominent networks (Figure 4; Figure S3). Next, given that multiple imagery conditions (i.e., appetitive, stress, neutral-relaxing) were combined in the predictive model, the contribution of each imagery condition was tested. All task conditions contributed to predicting craving (appetitive=29.5%, stress=27.0%, neutral-relaxing=43.5%) and the distribution of task contributions did not significantly differ from a uniform distribution (χ^2^=0.05, p=0.80, df=2). For the appetitive condition, nodes with the largest contribution to craving prediction were located in the PCC and primary sensory areas, with the motor-sensory network and DMN as the top contributing networks. Similarly, for the neutral-relaxing condition, nodes with the largest contribution to craving prediction were located in the PCC and primary sensory areas, plus the hippocampus and visual cortex, with the visual networks contributing most to prediction at the network level. In contrast, for the stress condition, nodes with the largest contribution to craving prediction were located in the parahippocampus and brain stem, as well as the PCC. Accordingly, the subcortical network (which includes the brain stem nodes) and the motor-sensory network were the top contributing networks. Finally, node and network contributions for the appetitive condition were significantly correlated with node and network contributions for both the stress (node: r=0.34, p<0.001, df=247; network: r=0.47, p<0.001, df=52) and neutral-relaxing conditions (node: r=0.35, p<0.001, df=247; network: r=0.31, p=0.02, df=52). However, node and network contributions for the stress and neutral-relaxing conditions were not correlated (node: r=-0.03, p=0.63, df=247; network: r=-0.01, p=0.95, df=55). Overall, model features were similar when excluding adolescent participants (r=0.53, p<0.001, df=247).

**Figure 3.**
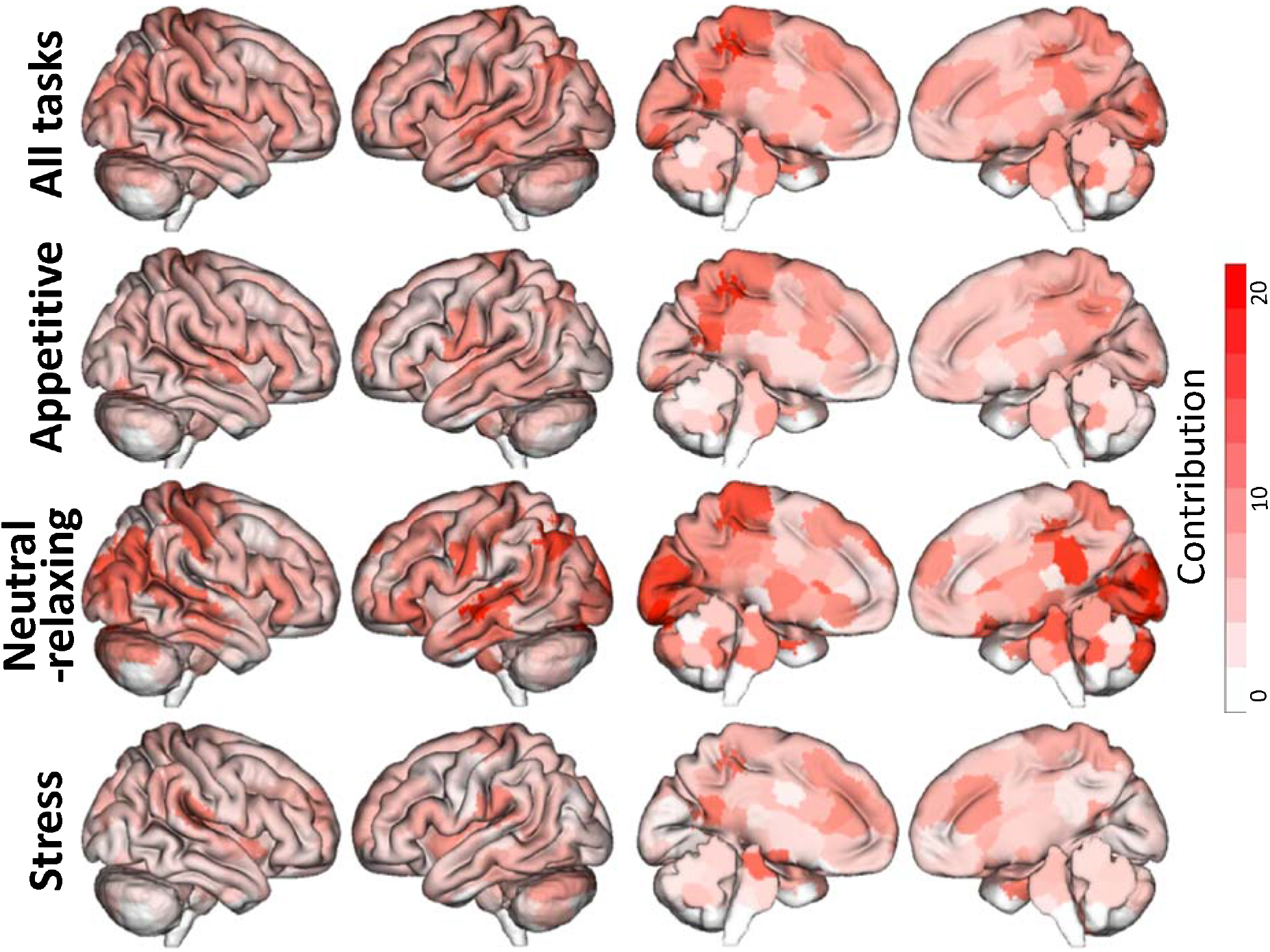
Node-level contribution to predictive craving values. The top row shows contribution (defined as the sum of all edgewise ridge regression coefficients normalized by the standard deviation of edges across participants for any node) across all three imagery conditions. The next three rows exhibit these contributions separated by each imagery condition. While all conditions highlight the PCC as a top contributor, the appetitive and neutral-relaxing conditions also highlighted contributions from primary sensory and visual networks as well as the hippocampus. The stress condition highlights regions in the brain stem and parahippocampus. Warmer colors represent nodes that contributed more towards the final prediction.

**Figure 4.**
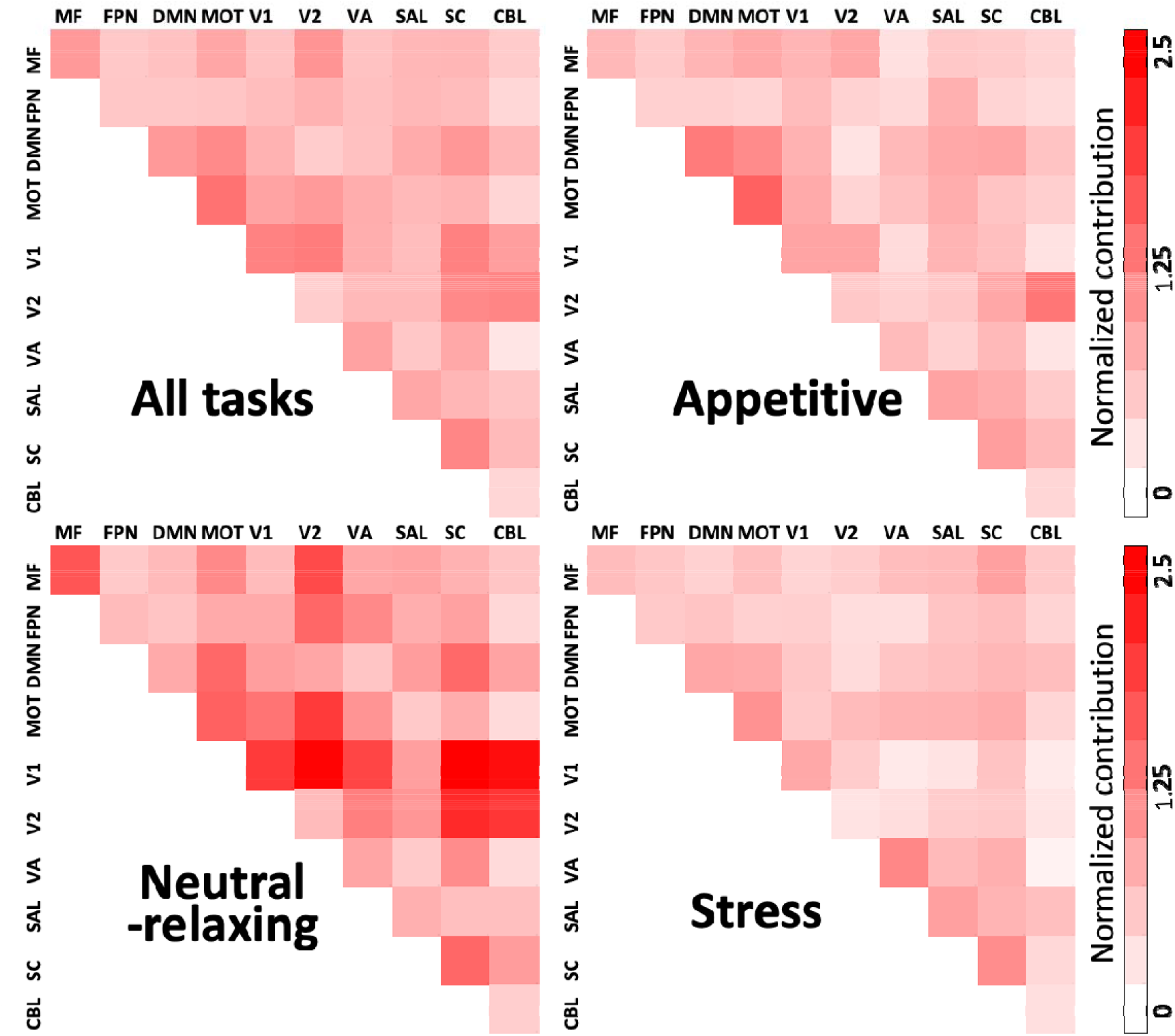
Network-level contribution to predictive craving values. Node-level contributions to predictions were further summarized by canonical functional networks. The diagonal represents the average contribution of edges within a single network, while off diagonal elements represent the average contribution of edges between two network pairs. Warmer colors represent networks that contributed more towards the final prediction. MF=medial frontal network; FPN=frontal parietal network; MOT=motor-sensory network; V1=visual network #1; V2=visual network #2; VA=visual association network; SAL=salience network; SC=Subcortical network; CBL=cerebellar network.

### Gender-related differences in predictions of craving

Based on putative sex/gender differences in the neural substrates of craving (e.g., ^31^), we investigated whether gender moderated the prediction of craving. The CPM analysis was repeated in each gender independently. Prediction performance was numerically higher but not significantly different in women compared to men (women: r=0.38, ρ=0.35, q^2^=0.13, MSE=34.93, n=178; men: r=0.20, ρ=0.20, q^2^=0.04, MSE=38.00, n=96; z=1.54, p=0.12). Likewise, models trained only in women predicted craving in men (r=0.39, p<0.001, df=176) and vice versa (r=0.43, p<0.001, df=94).

### Group differences in connectivity

Finally, we investigated group differences in task connectomes (i.e., appetitive, stress, neutral-relaxing connectomes) between the substance-use-related and control groups and how these group differences overlapped with the identified ‘craving network’.

Across all conditions, 1547 edges, or about 5% of the total number of possible edges, exhibited significant (p<0.05, corrected) group differences in connectivity between the substance-use-related and control groups (Figure 5). The nodes with the largest number of significant edges that differed between groups were located in the cerebellum, brainstem, PCC, and frontoparietal association cortices. When considering these edges across conditions, stronger connectivity during the appetitive condition, and both stronger and weaker connectivity during the stress condition (i.e., for different edges), for the substance-use-related group compared to control group, contributed most of these edges (691, 421, 410, respectively). In general, the group differences in cerebellar and brainstem edges were observed during the appetitive condition, while the group differences in the PCC and association cortices were observed during the stress condition. No significant group differences during the neutral-relaxing condition were observed.

**Figure 5.**
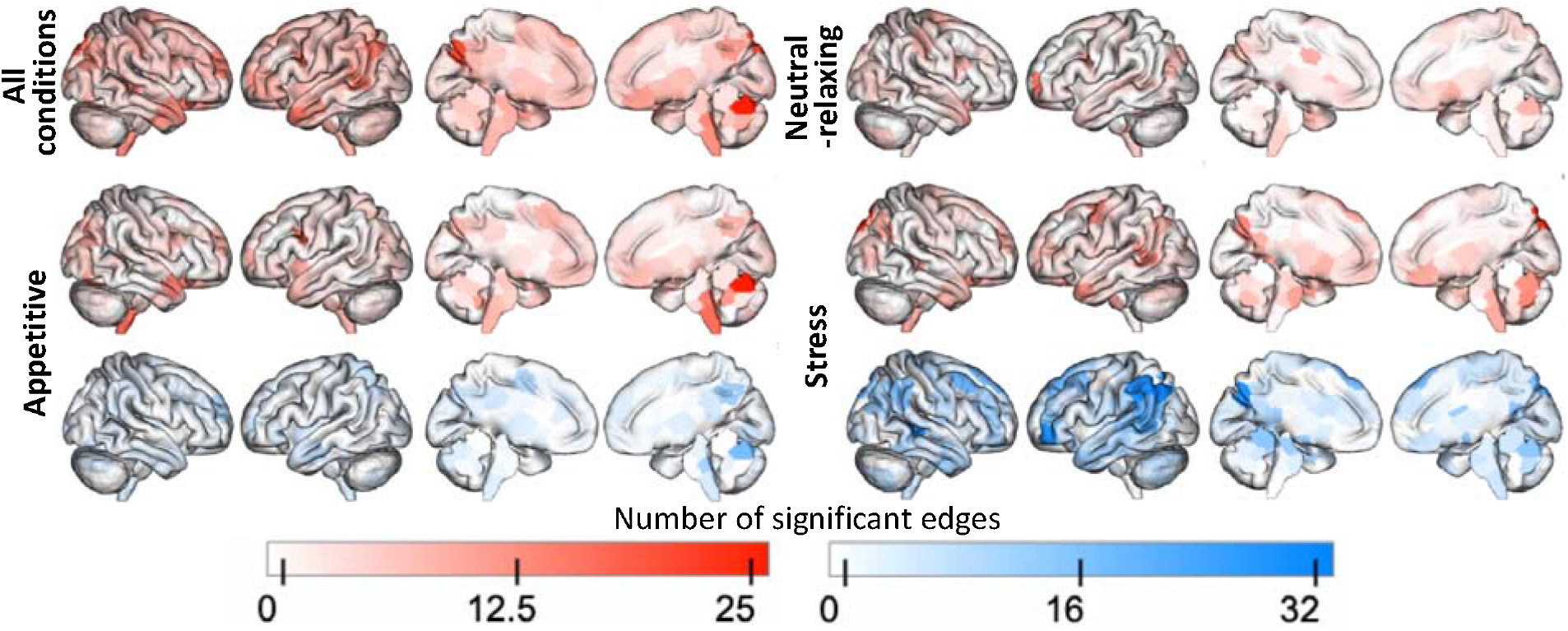
Node-level contribution to group differences in connectivity. One-thousand-five-hundred-and-forty-seven edges, or about 5% of the total number of possible edges, exhibited significant (p<0.05, corrected) group differences in connectivity between the substance use-related and control groups. The nodes with the largest number of significant edges that differed between groups were located in the cerebellum, brainstem, PCC, and frontoparietal association cortices. Overall, the nodes contributing to prediction and the nodes contributing to the group differences were different and not overlapping.

Overall, the nodes contributing to prediction and the nodes contributing to group differences were not correlated, suggesting that the predictive modeling results are neurobiologically distinct from the group differences (all conditions: r=-0.07, p=0.25, df=245; appetitive: r=-0.021, p=0.75, df=245; stress: r=0.01, p=0.92, df=245; neutral-relaxing: r=-0.021, p=0.75, df=245).

## Discussion

In this study, CPM was used to identify a neurofunctional ‘craving network’ in a large, transdiagnostic sample of individuals with and without substance-use-related conditions. Functional connectivity in the identified ‘craving network’ during an imagery task predicted a continuous measure of self-reported craving across individuals with alcohol or cocaine use disorders, obesity, adolescents with and without prenatal cocaine exposure, and adult controls. Importantly, the neuroanatomical nodes contributing to prediction and group differences were not correlated, suggesting the ‘craving network’ is neurobiologically distinct from group differences. These data provide a transdiagnostic perspective to a key phenomenological feature of addictive disorders—craving—and identify a common ‘craving network’ across disordered and non-disordered individuals, thereby suggesting a neural signature for craving/urge for motivated behaviors.

Consistent with the CPM approach, the ‘craving network’ was found to be complex and distributed throughout the brain. Studies of psychiatric disorders more generally, ^32^ and of SUDs, ^33,34^ are increasingly focused on understanding how distributed brain regions organized as large-scale brain networks contribute to dysfunction. ^32^ The strongest predictor of craving in the current study was functional connectivity in regions of the DMN, a core functional network. ^35^ The DMN is implicated in internally-directed thought processes as opposed to externally-directed processing (i.e., to external stimuli). ^36^ Internally-directed thought processes include self-related cognitive activities, including autobiographical, self-monitoring and social processes. ^32^ The current finding is consistent with a large-scale brain network model in which a lack of suppression of the DMN results in altered self-referential processing, ^32^ such as difficulties disengaging from the internal experiences of craving. ^28,34^ This leads to the speculation that such engagement of the DMN represents urge and intent for motivated behavior in response to various appetitive and stress cues that represent key contexts that may drive motivated behavior.

Altered DMN processing has been identified across SUDs and is associated with craving and relapse. ^37^ Exposure to substance cues has been found to increase activity in the posterior DMN (i.e., the precuneus/PCC) (e.g., for cocaine and alcohol ^38-40^). A lack of suppression of activity in the posterior DMN during task performance has been found in individuals during nicotine abstinence ^41^ and was related to smoking relapse. ^42^ Similarly, resting state functional connectivity in the posterior DMN has been found to be greater during abstinence and associated with relapse across different SUDs (e.g., opioids, cannabis and alcohol: ^43-45^). In a case study, an individual with stroke and lesion to the PCC reported a complete disruption of smoking addiction, including 12-month abstinence and a lack of urges to smoke. ^46^ An interpretation of these findings, in agreement with the current findings, is that substance use, in particular the withdrawal and preoccupation phases of the addiction cycle, is associated with difficulties directing attention away from self-referential processing, such as craving, represented in the posterior DMN. DMN suppression has also been related to cognitive control in SUDs (e.g., cocaine ^47^). Together, the findings suggest that aberrant DMN suppression may contribute to multiple processes (craving, cognitive control) linked to SUDs.

It is notable that the anterior DMN (i.e., the medial prefrontal cortex [MPFC]) was not found to be predictive of craving. The anterior and posterior midline DMN have distinct roles in self-referential processing, with the anterior DMN implicated largely in the attribution of personal value, and the posterior DMN implicated in internally-directed attention. ^37^ For example, one study demonstrated that explicit self-reference engages the MPFC, rest engages the precuneus, and both self-reference and rest engage the PCC. ^48^ Furthermore, and in line with the fMRI task used in the current study, this distinction has been summarized as PCC involvement in retrieval of autobiographical episodic memory and self-referential tasks, and MPFC involvement in social cognitive tasks and self-referential tasks. ^49^ One interpretation is that craving in response to the imagery provocation task was related to internally-directed attention and autobiographical memory processes, represented in the posterior DMN, and less to emotional processing represented in the anterior DMN, in particular, given the large control sample contributing to the overall model. Related to this, functional connectivity in the hippocampus was also found to be predictive of craving in the current study. The hippocampus is considered part of the DMN involved in past and future autobiographical thought and episodic/contextual retrieval,^49^ such as constructing a mental scene based on memory. ^50^ Similarly, primary sensory areas were involved, likely related to sensory processes during mental imagery. ^51^

Mounting evidence suggests that alterations in the organization and function of the DMN are common features across psychiatric and neurologic disorders, ^32,49^ including SUDs. Transdiagnostic studies, such as the current study, can be used to relate heterogeneous phenotypes (i.e., wanting/urges/craving) to specific brain networks (i.e., posterior DMN), which can then be related to clinically relevant variations, and to genetic, molecular and cellular factors. ^52^ Such findings can also be used to evaluate how brain networks are uniquely altered in specific disorders. ^27^ The current findings suggest that increased DMN connectivity is a common feature of subjective craving, which may influence motivated behavior in response to rewarding or appetitive stimuli, across individuals with or without substance use-related conditions.

In this study, participants were grouped as individuals with and without substance-use-related conditions, with several subgroups (i.e., SUDs, obesity, prenatal cocaine exposure) included in the former. Likely important differences in brain networks related to craving exist between these subgroups. However, the purpose of this study was to identify a common ‘craving network’ across groups. Furthermore, each subgroup sample size was considered small for neuroimaging-based predictive modeling, where sample sizes are recommended to be >100-200, ^53^ therefore the study was underpowered to test subgroups independently and to identify differential components of craving across specific subgroups, which remains for future work. Group differences were identified overall between individuals with and without substance-use-related conditions, in the cerebellum, brainstem, PCC, and frontoparietal association cortices.

However, the nodes contributing to prediction and group differences were not correlated, suggesting the ‘craving network’ is neurobiologically distinct from group differences and may therefore relate to craving/wanting as a more central construct driving human motivated behavior. ^17^

The craving measure used here was a single item, self-reported craving rating, measured at multiple time points (i.e., before and after each imagery condition) across the fMRI study. Because craving might change with exposure to appetitive, stress and neutral-relaxing imagery, prediction performance was compared for craving measured before and after imagery. While prediction accuracy was high for craving at both time points, prediction of craving measured before imagery was better than prediction of craving measured after imagery. It is possible that substance-use-related and control groups were more similar, in terms of brain-phenotype association, prior to imagery, leading to better prediction. After imagery, the similarities between groups, and therefore prediction accuracy, were reduced. As there was no time-by-group interaction, this suggests greater variance in connectivity data in the substance-use-related group as an explanation. This interpretation is supported by the group differences in the ‘craving network’ noted above. Nevertheless, prediction accuracy was high for craving measured before and after imagery, suggesting group differences are subtle, and lending support to the transdiagnostic model. This finding is also in line with evidence that craving is influenced by individual variation in craving responding and other contextual factors,^54^ with such variation contributing to better prediction in the model.

In this study, functional-connectivity matrices, or connectomes, across three imagery conditions (appetitive, stress, neutral-relaxing), were combined in a single model to predict craving. Task conditions were used because, compared with resting-state data, predictive modeling of task data has been found to better detect individual differences ^55,56^ and improve prediction of phenotypes. ^22,57^ Connectomes were combined across multiple task conditions because this approach has been found to outperform prediction from single task connectomes, ^29^ and further, the resulting predictive models should be more generalizable. However, different task connectomes have been found to contribute differently to final predictive models; ^29^ therefore, we tested the contribution of each imagery condition to the final predictive model of craving. Findings indicated that all three imagery conditions contributed to predicting craving, and the combined model outperformed prediction using any single condition and craving. As studies have found that task conditions are better at generating models of phenotypes related to the circuits they perturb, ^22,57^ future studies might test predictive models of craving using alternate task data, such as visual substance cue-reactivity.

Findings indicated that prediction performance did not differ significantly between genders, and models trained only in women predicted craving in men, and vice versa. Prediction performance was found to be numerically higher in women, possibly due to the nearly 2:1 ratio of women to men in the sample. Future studies might test how the ‘craving network’ differs by gender, such as related to clinical outcomes in substance-use-related disorders.

This study had some limitations. The craving measure used was self-reported, and was therefore sensitive, as with all subjective measures, to response bias, introspective abilities/tendencies and other factors. Some machine-learning studies have found higher prediction accuracy for objective measures than for subjective ratings (e.g. ^58^). However, there is no single accepted measure of craving, and nearly all models of craving assume that it can be measured, at least in part, by self-report. ^54,59^ The findings are also generalizable to much of the research on substance craving, which uses self-report. ^54^ Furthermore, craving was measured in the moment, at multiple time points and was highly correlated across time and conditions, thereby reducing potential noise. It would be interesting to test the resulting predictive model related to objective measures of craving (e.g., physiological, behavioral) in future studies, although self-reported craving does relate to intake behavior of food ^4^ and drugs. ^11^ In addition, the functional significance of the ‘craving network’ to other aspects of substance use or addictive behaviors has yet to be determined. While some potential confounds were controlled, there may yet be effects of others, including clinical and cognitive variables such as substance-use severity and cognitive impairments.

In conclusion, this study used CPM to identify a transdiagnostic ‘craving network’ in individuals with and without substance use-related conditions. The strongest predictor of craving was functional connectivity in regions of the DMN, consistent with a role for the DMN in self-referential processing and internally-directed attention, shown to be altered in individuals with SUDs and related to craving. These findings have several potential clinical applications. The DMN has been suggested as a potential biomarker for substance-use risk and treatment response, and a treatment target for SUDs. ^37^ The identified ‘craving network’ might be utilized similarly, in particular for individuals who report high levels of craving; ^60^ for treatments that target craving, such as medication, ^61^ behavioral therapies including cognitive behavioral therapy, ^62^ mindfulness-based interventions, ^63-65^ and substance-use exposure; ^66^ and for novel, brain-based interventions such as neurofeedback targeting craving ^67,68^ and/or the DMN, ^69,70^ or targeting the ‘craving network’ directly. ^71^

## Methods

### Data source

Data were included from several prior studies ^31,72-76^ utilizing a personalized guided imagery task during fMRI. All participants provided written informed consent for those studies, approved by the Yale School of Medicine Human Investigation Committee (Yale’s Institutional Review Board). Participants were included in the current analysis if they had complete data for all three fMRI imagery conditions, had complete data for self-reported craving, and had an average head motion less than 0.2 mm across all fMRI conditions.

### Participants

Participants (N=274) included individuals with alcohol use disorder (n=38), cocaine use disorder (n=28), obesity (body mass index >30 kg/m^2^; n=19), and adolescents with prenatal cocaine exposure (n=41), as well as controls (n=148) which included adult controls (n=131) and adolescents without prenatal cocaine exposure (n=17). Note that adult controls included a subgroup of social drinkers used for comparison in the earlier study of alcohol use disorder. Participants were median age 23 years (range 14-45 years), with 96 men and 178 women. Of the sample, 52% were White, 37% were Black or African American, 4% were Asian, 4% were biracial, 1% were other race, and 2% were unknown race. Participants with alcohol or cocaine use disorder had a current diagnosis of alcohol or cocaine dependence as determined by a Structured Clinical Interview for DSM-IV, self-reported alcohol or cocaine use prior to admission, and verified by urine toxicology. No other study participants met criteria for a current Axis I disorder including SUDs. All participants were free of significant neurological or psychiatric disorders or medical conditions. Additional exclusion criteria included inability to read and write in English, use of psychotropic medications, history of head trauma, pregnancy, claustrophobia or metal in body incompatible with MRI. For more detailed participant characteristics, see the original reports. ^31,72-76^

### Imagery paradigm

During fMRI, participants were presented with six personalized guided imagery scripts: two appetitive, two stress and two neutral-relaxing scripts. For the appetitive condition, in the substance use-related group, individuals with alcohol use disorder were presented with alcohol cue scripts, those with cocaine use disorder were presented with cocaine cue scripts, and those with obesity or prenatal cocaine exposure were presented with favorite-food cue scripts. In the control group, adult social drinkers were presented with alcohol cue scripts, and all other controls were presented with favorite-food cue scripts. Standardized structured interviews were conducted using the Scene Development Questionnaire ^77^ to develop personally tailored scripts. ^77-79^ Appetitive scripts were based on participants’ experiences of substance (i.e., drug, alcohol, food) anticipation and consumption (e.g., birthday celebration, meeting friends at a bar; pizza, ice cream). Stress scripts were based on their experiences that made them “sad, mad or upset in that moment and they could do nothing to change it” (e.g., death in the family, romantic break-up). Appetitive and stress scripts were calibrated to ensure equivalent emotional valence across participants. Neutral-relaxing scripts were based on their experiences of neutral or relaxing situations (e.g., sitting in the park).

Imagery scripts were presented in a blocked design with 5.5 minute blocks, each comprised of a 1.5 minute quiet baseline, followed by a 2.5 minute imagery script, and a 1 minute quiet recovery. During baseline, participants were instructed to lay still and not do anything. During the recovery, participants were instructed to stop imagining and lay still for another minute. The order of the imagery conditions was randomized and counterbalanced, with no condition repeated consecutively, and each script presented only once. Following each block, participants took part in 2 minute of progressive relaxation, where they were instructed to relax muscles in each part of the body (i.e., to relax physiological muscle tension rather than mental relaxation or imagery).

Before and after each imagery script, participants rated their craving and anxiety on verbal analog scales from 1 anchored at “not at all” to 10 for “more than ever” or “extremely high.” Craving ratings indicated their “desire to [use substance] at that moment.” Anxiety ratings indicated how “tense, anxious and/or jittery” they felt. Heart rate was also monitored during fMRI using a pulse oximeter. Each subsequent imagery block began only once participants’ heart rate and subjective ratings had returned to baseline.

### Predictive modeling framework

For connectivity processing and construction of connectomes, see supplement. For predictive modeling, a multidimensional approach was employed, which combines multiple connectomes and behavioral measures into a single predictive modeling framework. ^25^ Given that a single behavioral measurement represents a noisy approximation of any behavioral construct, and there were six craving ratings (i.e., before and after each imagery script), a principal components analysis (PCA) was used to summarize craving in a data-driven manner for predictive modeling with CPM. To maintain separate train and test groups, for each iteration, each PCA was limited to the training datasets and the PCA coefficients applied to the test dataset. To generate predictive models of craving, all task connectomes were combined into a single predictive model using ridge regression. ^29^ Using multiple connectomes improves predictive modeling and facilitates a more holistic characterization of brain-behavior associations. ^29,30^ However, there is a high degree of similarity between connectomes from different tasks and edges from these connectomes are not independent. In brief, ridge regression accounts for these dependencies in a principled manner. For feature selection, a significance threshold of p<0.05 was used to select edges that are positively and negatively associated with craving across individuals in the training data. Various potential confounds (head motion, age, group, gender, anxiety, heart rate, smoking status) were controlled for at this feature selection step using partial correlation. ^80^

### Model validation

Two validation approaches were used. First, 10-fold cross-validation was used to train transdiagnostic models of craving. For 10-fold cross-validation, the total sample (N=274) was randomly divided—regardless of diagnostic group—into 10 approximately equal-sized groups; on each fold, the model was trained on 9 groups and tested on the excluded 10^th^ group. This procedure was repeated for 100 random divisions. Second, leave-one-group-out cross validation was used to test whether prediction results were driven by group differences in prediction performance from including both controls and individuals with alcohol or cocaine use disorder, obesity, or adolescents with prenatal cocaine exposure in a single model. In this analysis, models were trained solely on the control group and tested in the substance-use-related group as well as trained in the substance-use-related group and tested in the control group.

### Assessing prediction performance

For the 10-fold cross-validation analyses, model performance was evaluated with a cross-validated coefficient of determination, labeled 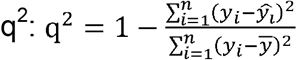, where *y*_*i*_ is the *i*^*th*^ observed value, *ŷ*_*i*_ is the *i*^*th*^ predicted value, and 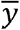 is the average of observed values. In the text, the median q^2^ for 100 random 10-fold divisions is reported. For convenience, Pearson’s correlation (r), Spearman’s rank correlation (ρ), and mean square error (MSE; defined as: 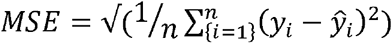 are also reported. To assess significance of q^2^, permutation testing was used, where the correspondence between behavioral variables and connectivity matrices were randomly shuffled 1,000 times and the CPM analysis was re-run with the shuffled data to generate null distributions of 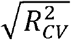. Based on these null distributions, the p-values for predictions were calculated as: p=(#[ρ_null>ρ_median]+1)/1001, where #[ρ_null>ρ_median] indicates the number of permutated predictions numerically greater than the median of the un-permutated predictions. As a positive association was expected between predicted and actual values, one-tailed p-values are reported. For the leave-one-group-out analysis, as it does not involve randomly partitioning data into training and testing sets, Pearson’s correlation was tested between actual and predicted craving to measure prediction performance. For quantification of task and anatomical contributions to prediction, see supplement.

### Group differences in connectomes

To measure group differences in task connectomes (i.e., appetitive, stress, neutral-relaxing connectomes) between the substance-use-related and control groups, mass univariate, edge-wise analyses was used, and multiple comparisons were controlled using the Network-Based Statistic (NBS). ^81^ For each edge, connectivity strength from each task-based connectome is included in a two-tailed t-test resulting in a node-by-node matrix of t-values that represent the magnitude of differences across groups. This procedure was performed independently for each task-based connectome. One-thousand iterations and an initial t-value threshold of 2.595 were used for NBS. Results at p<0.0167 (0.05/3) were considered significant. For the comparisons with the predictive modeling results, node contributions were quantified at the sum of all significant edges for a node. Edges showing greater and weaker connectivity between groups were summarized independently. In addition, results were also summarized across all task connectomes by creating the union of all significant edges from each task connectome, and summarizing node contribution as above.

### Model overlap

To assess the overlap between the transdiagnostic, predictive model of craving and group differences in connectomes, the node-level contributions were correlated between predictive models and group difference results.

### Code and model availability

Matlab scripts to run the main CPM analyses can be found at https://github.com/YaleMRRC/CPM/tree/master/matlab/func/misc. BioImage Suite tools used for analysis and visualization can be accessed at www.bisweb.yale.edu.

## Supporting information

see supplement

## Acknowledgements

Mind and Life Institute A-36649338, R01-AA013892-15, P50-DA016556, UL1-De019586, PL1-DA024859, RL1-AA017539, R01-DK00039, R01 MH121095, R01 DA039136

## Author Contributions

RS and MNP designed and conducted the original imagery experiments. CL contributed to the original data analysis and provided data management. KAG and DS conceived of the current study. DS led the analysis. DS, SG and QL conducted the analysis. KAG and DS wrote the paper. KAG, DS, RS and MNP discussed the results and implications and edited the paper.

## References

1 Kozlowski, L. T., Mann, R. E., Wilkinson, D. A. & Poulos, C. X. “Cravings” are ambiguous: Ask about urges or desires. Addict. Behav. 14, 443–445, doi:10.1016/0306-4603(89)90031-2 (1989).

2 Drummond, D. C. Theories of drug craving, ancient and modern. Addiction 96, 33–46, doi:10.1080/09652140020016941 (2001).

3 Carter, B. L. & Tiffany, S. T. Meta-analysis of cue-reactivity in addiction research. Addiction 94, 327–340 (1999).

4 Boswell, R. G. & Kober, H. Food cue reactivity and craving predict eating and weight gain: a meta-analytic review. Obes Rev 17, 159–177, doi:10.1111/obr.12354 (2016).

5 Kim, H. S., Hodgins, D. C., Kim, B. & Wild, T. C. Transdiagnostic or Disorder Specific? Indicators of Substance and Behavioral Addictions Nominated by People with Lived Experience. J Clin Med 9, doi:10.3390/jcm9020334 (2020).

6 Sinha, R. et al. Effects of adrenal sensitivity, stress-and cue-induced craving, and anxiety on subsequent alcohol relapse and treatment outcomes. Arch. Gen. Psychiatry 68, 942–952, doi:10.1001/archgenpsychiatry.2011.49 (2011).

7 Rohsenow, D. J., Martin, R. A., Eaton, C. A. & Monti, P. M. Cocaine craving as a predictor of treatment attrition and outcomes after residential treatment for cocaine dependence. J Stud Alcohol Drugs 68, 641–648, doi:10.15288/jsad.2007.68.641 (2007).

8 Waters, A. J. et al. Cue-provoked craving and nicotine replacement therapy in smoking cessation. J. Consult. Clin. Psychol. 72, 1136–1143, doi:10.1037/0022-006X.72.6.1136 (2004).

9 Moore, T. M. et al. Ecological momentary assessment of the effects of craving and affect on risk for relapse during substance abuse treatment. Psychol Addict Behav 28, 619–624, doi:10.1037/a0034127 (2014).

10 Yucel, M. et al. A transdiagnostic dimensional approach towards a neuropsychological assessment for addiction: an international Delphi consensus study. Addiction 114, 1095–1109, doi:10.1111/add.14424 (2019).

11 Tiffany, S. T. & Wray, J. M. The clinical significance of drug craving. Ann. N. Y. Acad. Sci. 1248, 1–17, doi:10.1111/j.1749-6632.2011.06298.x (2012).

12 Center for Behavioral Health Statistics and Quality. Impact of the DSM-IV to DSM-5 Changes on the National Survey on Drug Use and Health., (Rockville, MD, 2016).

13 Hasin, D. S., Fenton, M. C., Beseler, C., Park, J. Y. & Wall, M. M. Analyses related to the development of DSM-5 criteria for substance use related disorders: 2. Proposed DSM-5 criteria for alcohol, cannabis, cocaine and heroin disorders in 663 substance abuse patients. Drug Alcohol Depend. 122, 28–37, doi:10.1016/j.drugalcdep.2011.09.005 (2012).

14 Chase, H. W., Eickhoff, S. B., Laird, A. R. & Hogarth, L. The neural basis of drug stimulus processing and craving: an activation likelihood estimation meta-analysis. Biol. Psychiatry 70, 785–793, doi:10.1016/j.biopsych.2011.05.025 (2011).

15 Kuhn, S. & Gallinat, J. Common biology of craving across legal and illegal drugs - a quantitative meta-analysis of cue-reactivity brain response. Eur. J. Neurosci. 33, 1318–1326, doi:10.1111/j.1460-9568.2010.07590.x (2011).

16 Rosenberg, M. D., Finn, E. S., Scheinost, D., Constable, R. T. & Chun, M. M. Characterizing Attention with Predictive Network Models. Trends Cogn Sci 21, 290–302, doi:10.1016/j.tics.2017.01.011 (2017).

17 Robinson, T. E. & Berridge, K. C. The neural basis of drug craving: An incentive-sensitization theory of addiction. Brain Research Reviews 18, 247–291 (1993).

18 Shen, X. et al. Using connectome-based predictive modeling to predict individual behavior from brain connectivity. Nat Protoc 12, 506–518, doi:10.1038/nprot.2016.178 (2017).

19 Yarkoni, T. & Westfall, J. Choosing Prediction Over Explanation in Psychology: Lessons From Machine Learning. Perspect Psychol Sci 12, 1100–1122, doi:10.1177/1745691617693393 (2017).

20 Whelan, R. & Garavan, H. When optimism hurts: inflated predictions in psychiatric neuroimaging. Biol Psychiatry 75, 746–748, doi:10.1016/j.biopsych.2013.05.014 (2014).

21 Finn, E. S. et al. Functional connectome fingerprinting: identifying individuals using patterns of brain connectivity. Nat. Neurosci. 18, 1664–1671, doi:10.1038/nn.4135 (2015).

22 Rosenberg, M. D. et al. A neuromarker of sustained attention from whole-brain functional connectivity. Nat. Neurosci. 19, 165–171, doi:10.1038/nn.4179 (2016).

23 Dubois, J., Galdi, P., Han, Y., Paul, L. K. & Adolphs, R. Resting-state functional brain connectivity best predicts the personality dimension of openness to experience. Personal Neurosci 1, doi:10.1017/pen.2018.8 (2018).

24 Lake, E. M. R. et al. The Functional Brain Organization of an Individual Allows Prediction of Measures of Social Abilities Transdiagnostically in Autism and Attention-Deficit/Hyperactivity Disorder. Biol. Psychiatry 86, 315–326, doi:10.1016/j.biopsych.2019.02.019 (2019).

25 Barron, D. S. et al. Transdiagnostic, Connectome-Based Prediction of Memory Constructs Across Psychiatric Disorders. Cereb. Cortex, doi:10.1093/cercor/bhaa371 (2020).

26 Yip, S. W., Scheinost, D., Potenza, M. N. & Carroll, K. M. Connectome-Based Prediction of Cocaine Abstinence. Am J Psychiatry 176, 156–164, doi:10.1176/appi.ajp.2018.17101147 (2019).

27 Lichenstein, S. D., Scheinost, D., Potenza, M. N., Carroll, K. M. & Yip, S. W. Dissociable neural substrates of opioid and cocaine use identified via connectome-based modelling. Mol. Psychiatry, doi:10.1038/s41380-019-0586-y (2019).

28 Lerman, C. et al. Large-scale brain network coupling predicts acute nicotine abstinence effects on craving and cognitive function. JAMA psychiatry 71, 523–530, doi:10.1001/jamapsychiatry.2013.4091 (2014).

29 Gao, S., Greene, A. S., Constable, R. T. & Scheinost, D. Combining multiple connectomes improves predictive modeling of phenotypic measures. Neuroimage 201, 116038, doi:10.1016/j.neuroimage.2019.116038 (2019).

30 Jiang, R. et al. Task-induced brain connectivity promotes the detection of individual differences in brain-behavior relationships. NeuroImage 207, 116370, doi:https://doi.org/10.1016/j.neuroimage.2019.116370 (2020).

31 Potenza, M. N. et al. Neural correlates of stress-induced and cue-induced drug craving: influences of sex and cocaine dependence. Am J Psychiatry 169, 406–414, doi:10.1176/appi.ajp.2011.11020289 (2012).

32 Menon, V. Large-scale brain networks and psychopathology: a unifying triple network model. Trends Cogn Sci 15, 483–506, doi:10.1016/j.tics.2011.08.003 (2011).

33 Ersche, K. D. et al. Brain networks underlying vulnerability and resilience to drug addiction. Proc. Natl. Acad. Sci. U. S. A. 117, 15253–15261, doi:10.1073/pnas.2002509117 (2020).

34 Sutherland, M. T., McHugh, M. J., Pariyadath, V. & Stein, E. A. Resting state functional connectivity in addiction: Lessons learned and a road ahead. Neuroimage 62, 2281–2295, doi:10.1016/j.neuroimage.2012.01.117 (2012).

35 Smith, S. M. et al. Correspondence of the brain’s functional architecture during activation and rest. Proc. Natl. Acad. Sci. U. S. A. 106, 13040–13045, doi:10.1073/pnas.0905267106 (2009).

36 Buckner, R. L., Andrews-Hanna, J. R. & Schacter, D. L. in The year in cognitive neuroscience 2008 (eds Alan Kingstone & Michael B. Miller) 1–38 (Blackwell Publishing, 2008).

37 Zhang, R. & Volkow, N. D. Brain default-mode network dysfunction in addiction. Neuroimage 200, 313–331, doi:10.1016/j.neuroimage.2019.06.036 (2019).

38 Garavan, H. et al. Cue-induced cocaine craving: neuroanatomical specificity for drug users and drug stimuli. Am J Psychiatry 157, 1789–1798, doi:10.1176/appi.ajp.157.11.1789 (2000).

39 Vollstadt-Klein, S. et al. Effects of cue-exposure treatment on neural cue reactivity in alcohol dependence: a randomized trial. Biol. Psychiatry 69, 1060–1066, doi:10.1016/j.biopsych.2010.12.016 (2011).

40 Tomasi, D. et al. Overlapping patterns of brain activation to food and cocaine cues in cocaine abusers: association to striatal D2/D3 receptors. Hum. Brain Mapp. 36, 120–136, doi:10.1002/hbm.22617 (2015).

41 Falcone, M. et al. Age-related differences in working memory deficits during nicotine withdrawal. Addict Biol 19, 907–917, doi:10.1111/adb.12051 (2014).

42 Loughead, J. et al. Working memory-related neural activity predicts future smoking relapse. Neuropsychopharmacology 40, 1311–1320, doi:10.1038/npp.2014.318 (2015).

43 Li, W. et al. Dysfunctional Default Mode Network in Methadone Treated Patients Who Have a Higher Heroin Relapse Risk. Sci Rep 5, 15181, doi:10.1038/srep15181 (2015).

44 Osuch, E. A. et al. Depression, marijuana use and early-onset marijuana use conferred unique effects on neural connectivity and cognition. Acta Psychiatr. Scand. 134, 399–409, doi:10.1111/acps.12629 (2016).

45 Zhu, X., Cortes, C. R., Mathur, K., Tomasi, D. & Momenan, R. Model-free functional connectivity and impulsivity correlates of alcohol dependence: a resting-state study. Addict Biol 22, 206–217, doi:10.1111/adb.12272 (2017).

46 Jarraya, B. et al. Disruption of cigarette smoking addiction after posterior cingulate damage. J. Neurosurg. 113, 1219–1221, doi:10.3171/2010.6.JNS10346 (2010).

47 Worhunsky, P. D. et al. Multimodal investigation of dopamine D2/D3 receptors, default mode network suppression, and cognitive control in cocaine-use disorder. Neuropsychopharmacology 46, 316–324, doi:10.1038/s41386-020-00874-7 (2021).

48 Whitfield-Gabrieli, S. et al. Associations and dissociations between default and self-reference networks in the human brain. Neuroimage 55, 225–232, doi:10.1016/j.neuroimage.2010.11.048 (2011).

49 Whitfield-Gabrieli, S. & Ford, J. M. Default mode network activity and connectivity in psychopathology. Annu Rev Clin Psychol 8, 49–76, doi:10.1146/annurev-clinpsy-032511-143049 (2012).

50 Andrews-Hanna, J. R., Reidler, J. S., Sepulcre, J., Poulin, R. & Buckner, R. L. Functional-anatomic fractionation of the brain’s default network. Neuron 65, 550–562, doi:10.1016/j.neuron.2010.02.005 (2010).

51 Kosslyn, S. M., Ganis, G. & Thompson, W. L. Neural foundations of imagery. Nat. Rev. Neurosci. 2, 635–642, doi:10.1038/35090055 (2001).

52 Insel, T. et al. Research domain criteria (RDoC): toward a new classification framework for research on mental disorders. Am J Psychiatry 167, 748–751, doi:10.1176/appi.ajp.2010.09091379 (2010).

53 Sui, J., Jiang, R., Bustillo, J. & Calhoun, V. Neuroimaging-based Individualized Prediction of Cognition and Behavior for Mental Disorders and Health: Methods and Promises. Biol. Psychiatry 88, 818–828, doi:10.1016/j.biopsych.2020.02.016 (2020).

54 Sayette, M. A. The Role of Craving in Substance Use Disorders: Theoretical and Methodological Issues. Annu Rev Clin Psychol 12, 407–433, doi:10.1146/annurev-clinpsy-021815-093351 (2016).

55 Finn, E. S. et al. Can brain state be manipulated to emphasize individual differences in functional connectivity? Neuroimage 160, 140–151, doi:10.1016/j.neuroimage.2017.03.064 (2017).

56 Vanderwal, T. et al. Individual differences in functional connectivity during naturalistic viewing conditions. Neuroimage 157, 521–530, doi:10.1016/j.neuroimage.2017.06.027 (2017).

57 Greene, A., Gao, S., Scheinost, D. & Constable, R. Task-induced brain state manipulation improves prediction of individual traits. Nature Communications 9, 2807, doi:10.1038/s41467-018-04920-3 (2018).

58 Li, J. et al. Global signal regression strengthens association between resting-state functional connectivity and behavior. Neuroimage 196, 126–141, doi:10.1016/j.neuroimage.2019.04.016 (2019).

59 Sayette, M. A. et al. The measurement of drug craving. Addiction 95 Suppl 2, S189–210 (2000).

60 Roos, C. R., Brewer, J. A., O’Malley, S. S. & Garrison, K. A. Baseline Craving Strength as a Prognostic Marker of Benefit from Smartphone App-Based Mindfulness Training for Smoking Cessation. Mindfulness 10, 2165–2171, doi:10.1007/s12671-019-01188-6 (2019).

61 O’Brien, C. P. Anticraving medications for relapse prevention: a possible new class of psychoactive medications. Am J Psychiatry 162, 1423–1431, doi:10.1176/appi.ajp.162.8.1423 (2005).

62 Magill, M. et al. The search for mechanisms of cognitive behavioral therapy for alcohol or other drug use disorders: A systematic review. Behav. Res. Ther. 131, 103648, doi:10.1016/j.brat.2020.103648 (2020).

63 Garrison, K. A. et al. Craving to Quit: A Randomized Controlled Trial of Smartphone app-based Mindfulness Training for Smoking Cessation. Nicotine Tob Res, doi:10.1093/ntr/nty126 (2018).

64 Bowen, S. et al. Relative efficacy of mindfulness-based relapse prevention, standard relapse prevention, and treatment as usual for substance use disorders: a randomized clinical trial. JAMA psychiatry 71, 547–556, doi:10.1001/jamapsychiatry.2013.4546 (2014).

65 Janes, A. C. et al. Quitting starts in the brain: a randomized controlled trial of app-based mindfulness shows decreases in neural responses to smoking cues that predict reductions in smoking. Neuropsychopharmacology 44, 1631–1638, doi:10.1038/s41386-019-0403-y (2019).

66 Xue, Y. X. et al. A memory retrieval-extinction procedure to prevent drug craving and relapse. Science 336, 241–245, doi:10.1126/science.1215070 (2012).

67 Li, X. et al. Volitional reduction of anterior cingulate cortex activity produces decreased cue craving in smoking cessation: a preliminary real-time fMRI study. Addict Biol 18, 739–748, doi:10.1111/j.1369-1600.2012.00449.x (2013).

68 Kim, D. Y., Yoo, S. S., Tegethoff, M., Meinlschmidt, G. & Lee, J. H. The inclusion of functional connectivity information into fMRI-based neurofeedback improves its efficacy in the reduction of cigarette cravings. J. Cogn. Neurosci. 27, 1552–1572, doi:10.1162/jocn_a_00802 (2015).

69 Bauer, C. C. C. et al. Real-time fMRI neurofeedback reduces auditory hallucinations and modulates resting state connectivity of involved brain regions: Part 2: Default mode network-preliminary evidence. Psychiatry Res. 284, 112770, doi:10.1016/j.psychres.2020.112770 (2020).

70 Garrison, K. A. et al. Real-time fMRI links subjective experience with brain activity during focused attention. Neuroimage 81, 110–118, doi:10.1016/j.neuroimage.2013.05.030 (2013).

71 Scheinost, D. et al. Connectome-based neurofeedback: A pilot study to improve sustained attention. Neuroimage 212, 116684, doi:10.1016/j.neuroimage.2020.116684 (2020).

72 Hommer, R. E. et al. Neural correlates of stress and favorite-food cue exposure in adolescents: a functional magnetic resonance imaging study. Hum. Brain Mapp. 34, 2561–2573, doi:10.1002/hbm.22089 (2013).

73 Jastreboff, A. M. et al. Neural correlates of stress-and food cue-induced food craving in obesity: association with insulin levels. Diabetes Care 36, 394–402, doi:10.2337/dc12-1112 (2013).

74 Seo, D. et al. Sex differences in neural responses to stress and alcohol context cues. Hum. Brain Mapp. 32, 1998–2013, doi:10.1002/hbm.21165 (2011).

75 Seo, D. et al. Disrupted ventromedial prefrontal function, alcohol craving, and subsequent relapse risk. JAMA psychiatry 70, 727–739, doi:10.1001/jamapsychiatry.2013.762 (2013).

76 Yip, S. W., Lacadie, C. M., Sinha, R., Mayes, L. C. & Potenza, M. N. Prenatal cocaine exposure, illicit-substance use and stress and craving processes during adolescence. Drug Alcohol Depend. 158, 76–85, doi:10.1016/j.drugalcdep.2015.11.012 (2016).

77 Sinha, R. Modeling stress and drug craving in the laboratory: implications for addiction treatment development. Addict Biol 14, 84–98, doi:10.1111/j.1369-1600.2008.00134.x (2009).

78 Hommer, R. E. et al. Neural correlates of stress and favorite-food cue exposure in adolescents: A functional magnetic resonance imaging study. Hum Brain Mapp, doi:10.1002/hbm.22089 (2012).

79 Jastreboff, A. M. et al. Neural Correlates of Stress-and Food Cue–Induced Food Craving in Obesity: Association with insulin levels. Diabetes Care 36, 394–402, doi:10.2337/dc12-1112 (2013).

80 Hsu, W.-T., Rosenberg, M. D., Scheinost, D., Constable, R. T. & Chun, M. M. Resting-state functional connectivity predicts neuroticism and extraversion in novel individuals. Social Cognitive and Affective Neuroscience 13, 224–232, doi:10.1093/scan/nsy002 (2018).

81 Zalesky, A., Fornito, A. & Bullmore, E. T. Network-based statistic: identifying differences in brain networks. Neuroimage 53, 1197–1207, doi:10.1016/j.neuroimage.2010.06.041 (2010).

