## Supplementary material for "Transdiagnostic connectome-based prediction of craving": see supplement

**Supplementary Methods**

### Image acquisition

All images were collected on 3T Siemens Trio MRI scanners at the Yale Magnetic Resonance Research Center. Specific image acquisition parameters can be found in the original papers for these datasets.

### Connectivity processing

Functional images were motion corrected using SPM12. All further analyses were performed using BioImage Suite 36 unless otherwise specified. Several covariates of no interest were regressed from the data including linear and quadratic drifts, mean cerebral-spinal-fluid (CSF) signal, mean white-matter signal, and mean gray matter signal. For additional control of possible motion-related confounds, a 24-parameter motion model (including six rigid-body motion parameters, six temporal derivatives, and these terms squared) was regressed from the data. The data were temporally smoothed with a Gaussian filter (approximate cutoff frequency=0.12Hz).

### Construction of connectomes

Nodes were defined using the Shen 268-node brain atlas, which includes the cortex, subcortex, and cerebellum as described in prior CPM work. ^37^ Twenty-one nodes located in the brain stem, inferior cerebellum, temporal pole, and inferior orbital frontal lobe (see Figure S1) were removed from analysis due incomplete brain coverage. The atlas was warped from MNI space into single-subject space via series of linear and non-linear transformations. ^38^ Task connectivity was calculated on the basis of the ‘raw’ task time courses, with no regression of task-evoked activity, which emphasizes individual differences in connectivity. This involved computation of the mean time courses for each of the 268 nodes (i.e., averaging the time courses of all constituent voxels). Node-by-node pairwise correlations were computed, and Pearson correlation coefficients were Fisher z-transformed to yield symmetric 268x268 connectivity matrices, in which each element of the matrix represents the connectivity strength between two individual nodes (i.e., ‘edge’).

### Quantification of task and anatomical contribution to prediction

Predictive networks identified using CPM are complex and composed of multiple brain regions and networks. To quantify the contribution of each edge to a given predictive model, we calculated the $k^{th}$ edge’s weight for $m^{th}$ task (labeled $W_{k,m}$) to the model as: $W_{k,m}=\boldsymbol{B}(k,m){{abs(\beta}_{m}}^{k})std(\boldsymbol{E}_{k}(:,m))$, where $\boldsymbol{B}(k,m)$indexes whether the $k^{th}$ edge is selected from the $m^{th}$ task, $std(\boldsymbol{E}_{k}(:,m))$ represents the standard deviation of the $k^{th}$ edge in the $m^{th}$ task, and ${\beta_{m}}^{k}$represents the weight learned by CPM for the $k^{th}$ edge in the $m^{th}$ task. To quantify the contribution of each node to a given predictive model, we calculated the $n^{th}$ node’s weight summed across all tasks and edges (labeled $W_{n}$) to the model as:$W_{n}=\sum_{k=1}^{35,778} \sum_{m=1}^{3} W_{k,m}$, for all $k$ edges connected to the $n^{th}$ node. Next, for the network level, $W_{k,m}$ was averaged over each edge within or between canonical functional networks, based on the functional networks presented in (Noble et al., 2017). Finally, we quantified the contribution of each task as: $W_{n}=\sum_{k=1}^{35,778} W_{k,m}$. To increase interpretablility, $W_{m}$ are then normalized to have sum of 1, $\sum_{m=1}^{3} W_{m}=1$ , so that it represents each task’s proportional contribution in the model.

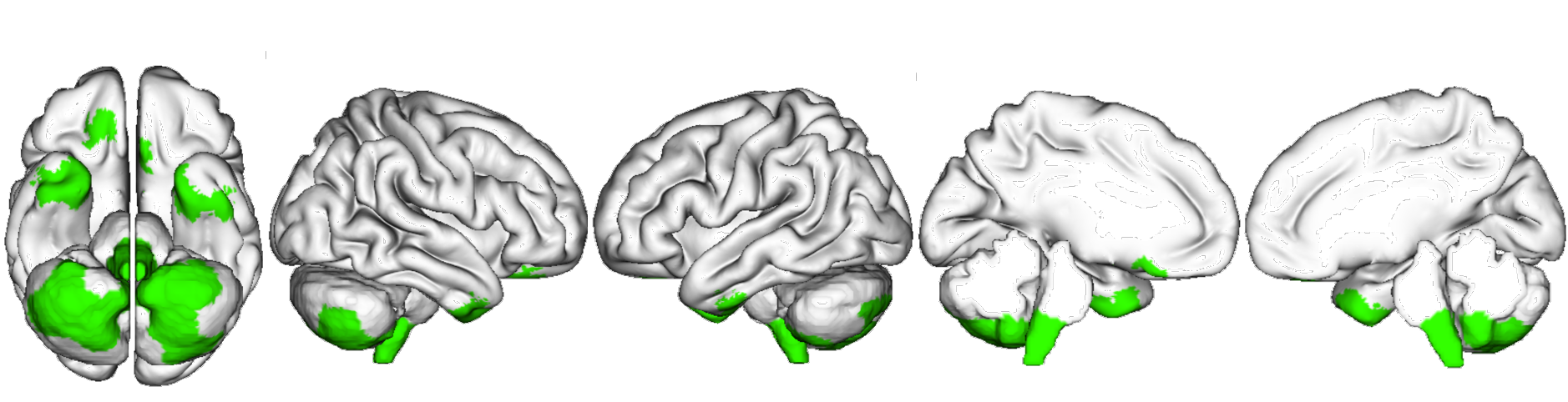

**Figure S1.** Spatial location of nodes removed from analysis due to incomplete slice coverage across subjects (green). These twenty-one nodes were located in the brain stem, inferior cerebellum, temporal pole, and inferior orbital frontal lobe.

**Supplementary Results**

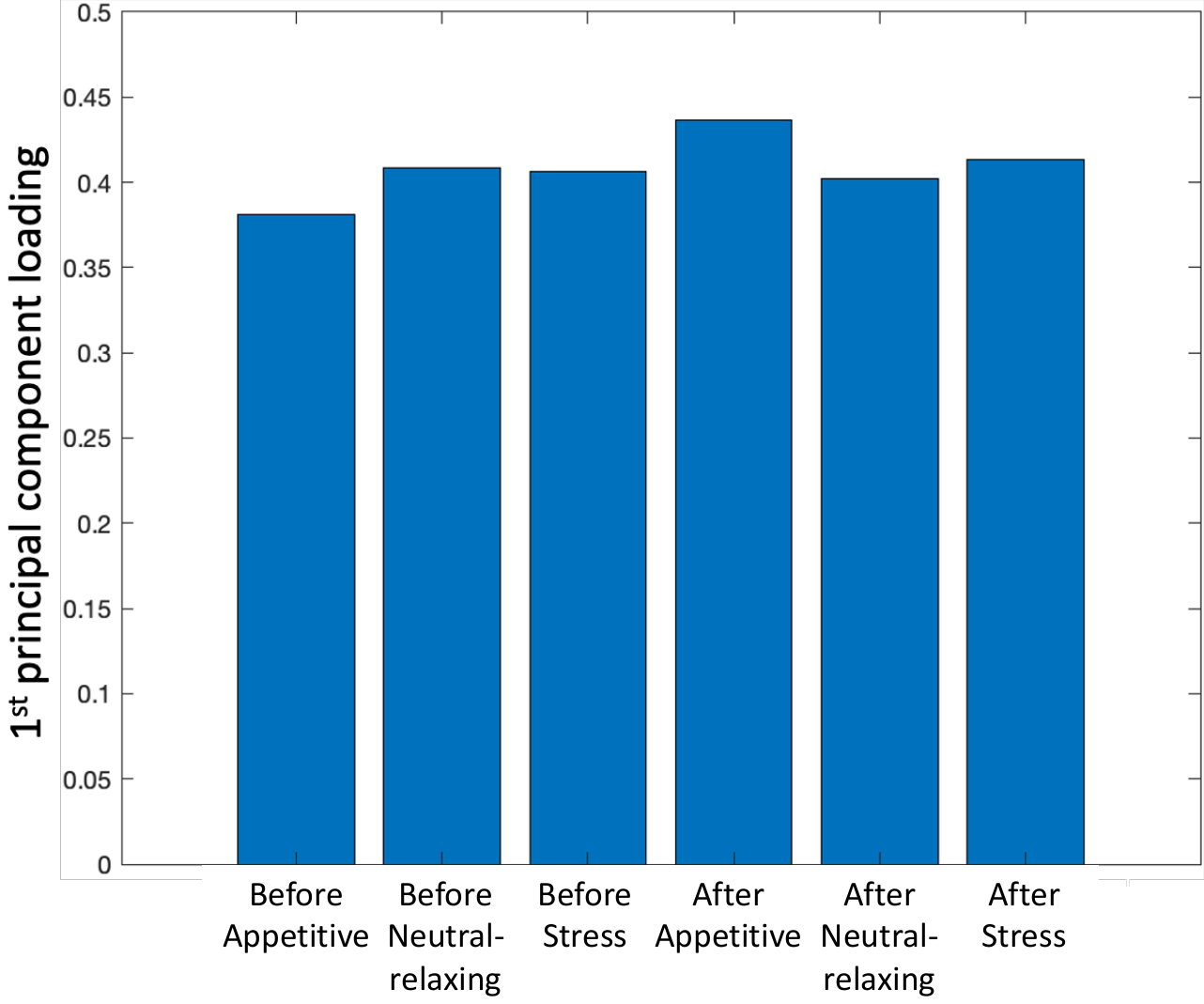

**Figure S2.** Loadings of the 1^st^ principal component across all craving measures. Across 6 self-report craving measures taken before and after each imagery condition, each craving measure contributed approximately equally to the 1^st^ principal component used in prediction.

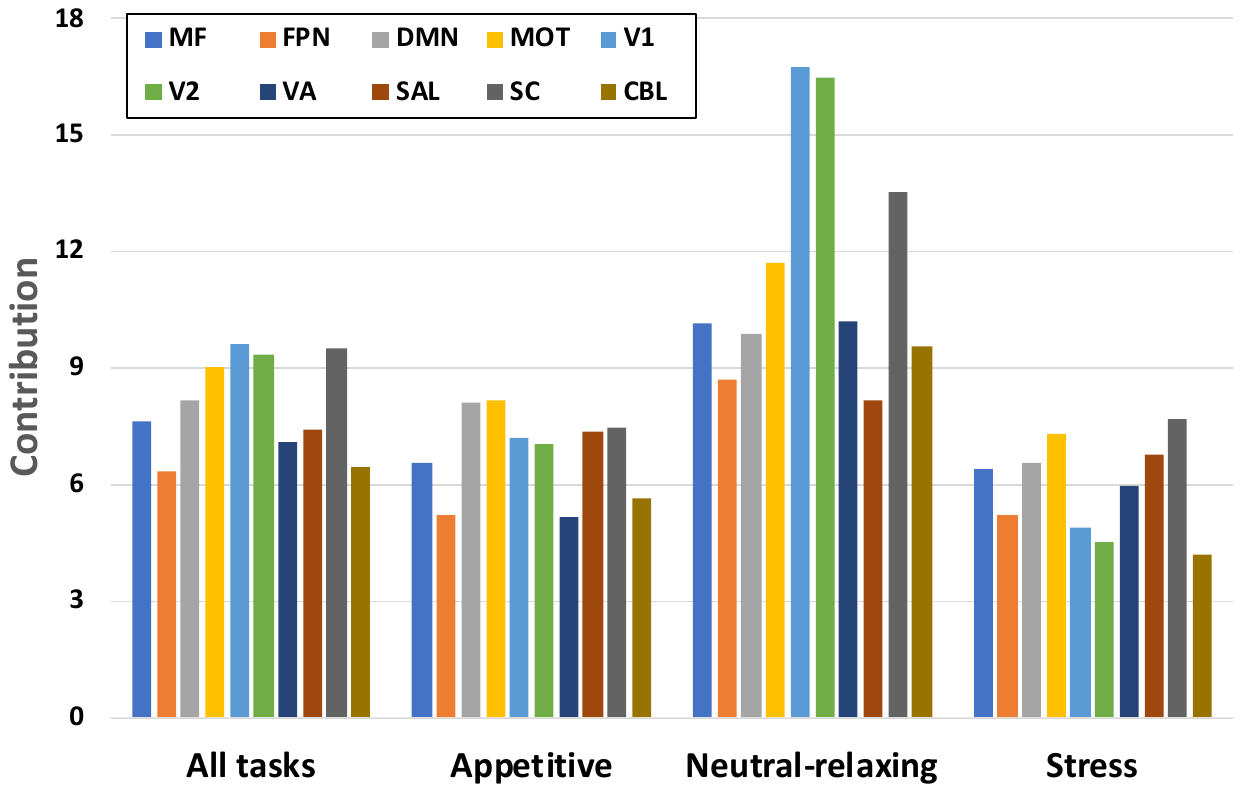

**Figure S3.** Total network-level contribution to predicted craving. Plotted values were calculated by sum over each each row (or column) in Figure 3 to produce a single summary number for total network-level contribution. For the appetive condition, the DMN and motor-sensory network contributed the most. For the neutral-relaxing condition, the visual networks contributed the most. For the stress condition, the subcortical and motor-sensory networks contributed the most. MF=medial frontal network; FPN=frontal parietal network; MOT=motor-sensory network; V1=visual network #1; V2=visual network #2; VA=visual association network; SAL=salience network; SC=subcortical network; CBL=cerebellar network.

### Table S1: Prediction perfromance using mean craving compared to PCA

| Variable | r | ρ | q^2^ | MSE | p-value |
| --- | --- | --- | --- | --- | --- |
| Mean craving across all conditions | 0.41 | 0.38 | 0.14 | 3.37 | p<0.001 |
| Mean craving before all conditions | 0.44 | 0.44 | 0.16 | 2.87 | p<0.001 |
| Mean craving after all conditions | 0.38 | 0.36 | 0.13 | 3.88 | p<0.001 |
| PCA on craving before all conditions | 0.45 | 0.45 | 0.17 | 16.37 | p<0.001 |
| PCA on craving after all conditions | 0.37 | 0.34 | 0.12 | 18.65 | p<0.001 |

Pearson’s correlation (r), Spearman’s rank correlation (ρ), cross-validated coefficient of determination (q^2^), and mean square error (MSE) between observed and predicted values

### Table S2: Prediction perfromance using a single imagery condition

| Task | r | ρ | q^2^ | MSE | p-value |
| --- | --- | --- | --- | --- | --- |
| Appetitive imagery alone | 0.29 | 0.28 | 0.06 | 3.57 | p<0.001 |
| Neutral-relaxing imagery alone | 0.36 | 0.35 | 0.1 | 2.78 | p<0.001 |
| Stress imagery alone | 0.29 | 0.34 | 0.05 | 3.07 | p<0.001 |

Pearson’s correlation (r), Spearman’s rank correlation (ρ), cross-validated coefficient of determination (q^2^), and mean square error (MSE) between observed and predicted values
